# Differential anchoring effects of vaccination comparator selection: characterizing a potential bias due to healthcare utilization in COVID-19 versus influenza

**DOI:** 10.1101/2021.10.07.21264711

**Authors:** Anna Ostropolets, Patrick B. Ryan, Martijn J. Schuemie, George Hripcsak

## Abstract

**Introduction:** Observational data enables large-scale vaccine safety surveillance but requires careful evaluation of potential sources of bias. One potential source of bias is an index date selection procedure for the unvaccinated cohort or unvaccinated comparison time. Here, we evaluate different index date selection procedures for two vaccines: COVID-19 and influenza.

**Methods:** For each vaccine, we extracted patient baseline characteristics on the index date and up to 450 days prior and then compared them to the characteristics of the unvaccinated patients indexed on an arbitrary date or indexed on a date of a visit. Additionally, we compared vaccinated patients indexed on the date of vaccination and the same patients indexed on a prior date or visit.

**Results:** COVID-19 vaccination and influenza vaccination differ drastically from each other in terms of populations vaccinated and their status on the day of vaccination. When compared to indexing on a visit in unvaccinated population, influenza vaccination had markedly higher covariate proportions and COVID-19 vaccination had lower proportions of most covariates on the index date. In contrast, COVID-19 vaccination had similar covariate proportions when compared to an arbitrary date. These effects attenuated but were still present with a longer lookback period. The effect of day 0 was present even when patients served as their own controls.

**Conclusion:** Patient baseline characteristics are sensitive to the choice of the index date. In vaccine safety studies, unexposed index event should represent vaccination settings. Study designs previously used to assess influenza vaccination must be reassessed for COVID-19 to account for a potentially healthier population and lack of medical activity on the day of vaccination.

## Introduction

The world is faced with a deadly pandemic at a time of incredible technology such that new vaccines can be produced in a fraction of the previous development time and at a scale that can potentially vaccinate the entire human population. This brings new challenges in using observational data to evaluate vaccine safety, where pressure to vaccinate quickly to prevent more deaths and more viral variants reduces time to carry out studies [1]. This time pressure affects not just the collection of data for research but even the time it takes to develop and validate evaluation methods. We therefore rely on methods developed and validated in previous pandemics and seasonal infectious diseases, with influenza being an important example [2–5].

COVID-19 vaccination has been unlike any other in history. The target vaccination group has shifted from the elderly and those with comorbidities in the early phases of vaccination to everyone including the healthy young [6], with some nations already vaccinating the majority of their populations [7]. COVID-19 vaccines are delivered in a wide variety of settings, from pop-up centers unconnected to health care delivery to inpatient facilities on hospital discharge. Other vaccines like those for influenza have a different delivery. They are often administered to specific vulnerable populations such as pregnant women, patients at high risk of complications or children, and are often given during health care visits [8–10].

The unique properties of COVID-19 vaccination may require adjusting study designs previously used for influenza vaccination, specifically the selection of a comparator cohort or an unvaccinated comparison time in cohort and self-controlled studies. While for the vaccinated group, the index date – vaccination - is clearly defined, selection of the index date for the unexposed comparator group is more complex. The selection procedure (which we have termed “anchoring”) may itself influence the results of a study and induce bias in the analysis. For example, in studies of background rates of adverse events, patients indexed on an arbitrary date were shown to have lower incidence of adverse events compared to the same patients indexed on a visit [11]. Ideally, the index date in the unexposed group should be chosen based on the vaccination settings to reliably serve as a counterfactual.

Here, we evaluate two alternative selection procedures based on how vaccines are administered - coupled or decoupled to another healthcare encounter. We compare these approaches for two vaccines: influenza and COVID-19.

## Methods

We studied two types of vaccination: influenza vaccine administered during 2017 – 2018 and COVID-19 vaccine administered during 2020 – 2021. For each vaccine, we mimicked two study designs. The first design (Figure 1, A) corresponds to a cohort method, where the target group was vaccinated patients, and the comparator group was unvaccinated patients. The index date for the target group was the date of vaccination; for the comparator it was (a) a date selected from the unvaccinated patient’s history (not necessarily with any medical event) such that it matched the index date of one of the target group participants or (b) a visit matched to the index date of one of the target group participants. Patients in each target and comparator pair were matched on age and sex. The second design corresponds to a self-controlled design (case-crossover design) [12] where the cases were the vaccinated patients indexed (or “anchored”) on the day of vaccination and the controls were the same patients indexed on an arbitrary date or a visit within 180-450 days prior to the vaccination date.

**Figure 1.**
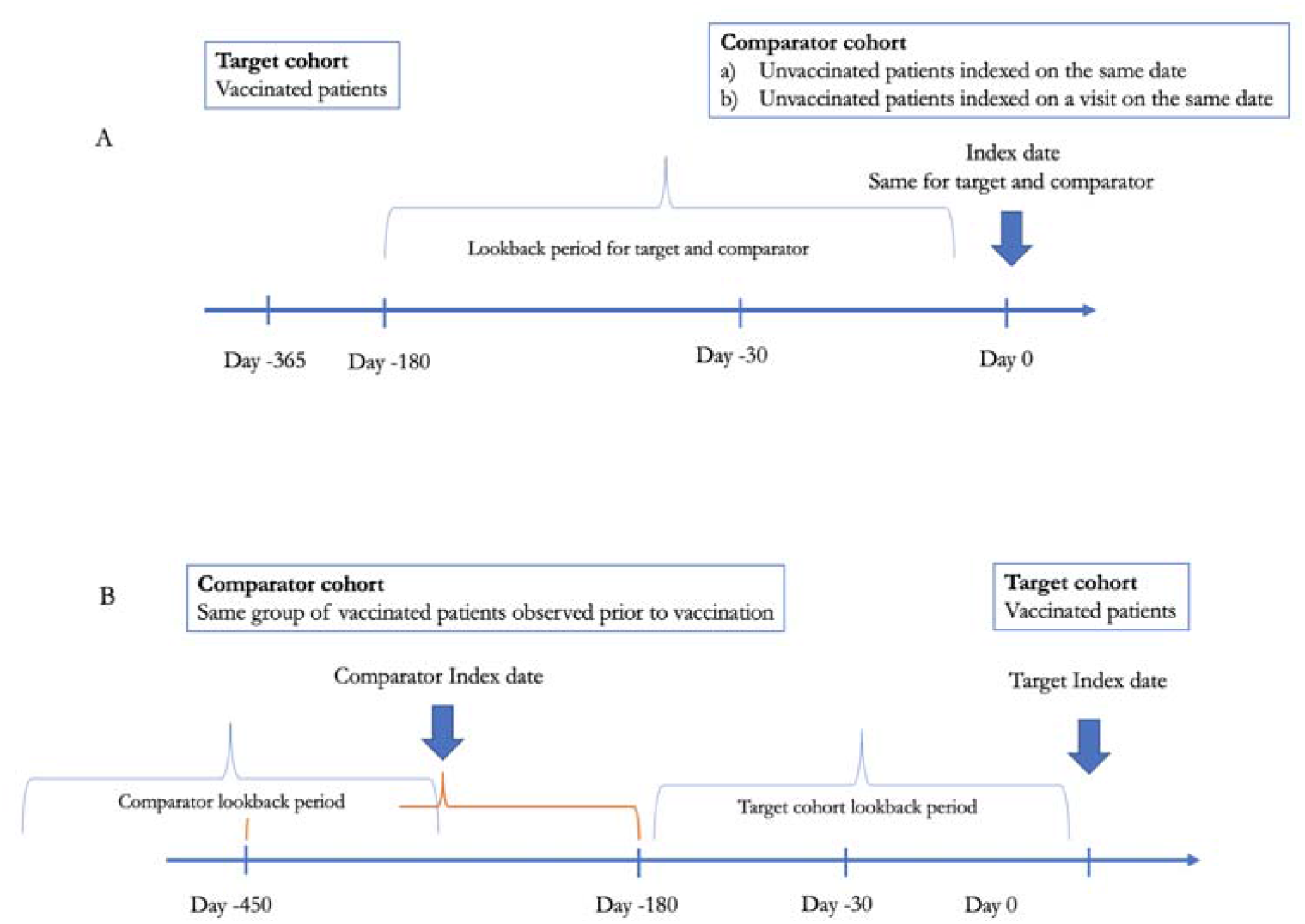
Study design overview

For each group, we collected patient baseline characteristics (covariates) recorded within five time intervals: at the index date (day 0), at the day before the index date (day -1), from 30 to 1 day prior to the index date (short term baseline covariates), from 180 to 31 day prior to the index date (medium term baseline covariates) and from 450 to 181 day prior to the index date (long term baseline covariates). Baseline covariates included all conditions, procedures, measurements and drug groups available in patient records within a specified time interval. For each covariate, we calculated covariate proportion, which is the proportion of patients with a covariate recorded in their electronic health record within a given time interval.

We conducted the analysis on two electronic health record data sources: Columbia University Medical Center Health Record Dataset (CUMC) and Optum© de-identified Electronic Health Record Dataset (Optum EHR). Optum EHR’s data comprises medical record data from 87 million patients and includes clinical information, inclusive of prescriptions as prescribed and administered, lab results, vital signs, body measurements, diagnoses, procedures, and information derived from clinical notes using natural language processing. CUMC EHR gathers data from the clinical data warehouse of NewYork-Presbyterian Hospital/Columbia University Irving Medical Center, New York, NY, based on its current and previous electronic health record systems, with data spanning over 30 years and including over 6 million patients. All data sources were mapped to the Observational Medical Outcomes Partnership Common Data Model (OMOP CDM) [13]. The OMOP CDM provides a homogeneous format for healthcare data and standardization of underlying clinical coding systems that thus enables analysis code to be shared across participating datasets in the network.

## Results

Comparison of vaccinated patients and unvaccinated patients indexed on a date or a visit

### Influenza vaccinated population

On the index date (day of vaccination = day 0), the influenza vaccinated population had markedly higher proportion of most covariates than an arbitrary date in the comparison group (pinning most covariates against the X-axis in Figure 2, A and B, yellow). The largest difference in covariate proportions between unvaccinated and vaccinated populations on day 0 was observed for inpatient and outpatient measurements such as blood count, metabolic panels, blood pressure and basal metabolic index, including both presence of measurements and proportion of patients with of abnormal results, meaning patients were far more likely to have measurements on the date of vaccination than on an arbitrary date. Moreover, influenza vaccinated population had higher covariate proportions even a year prior to the vaccination.

**Figure 2.**
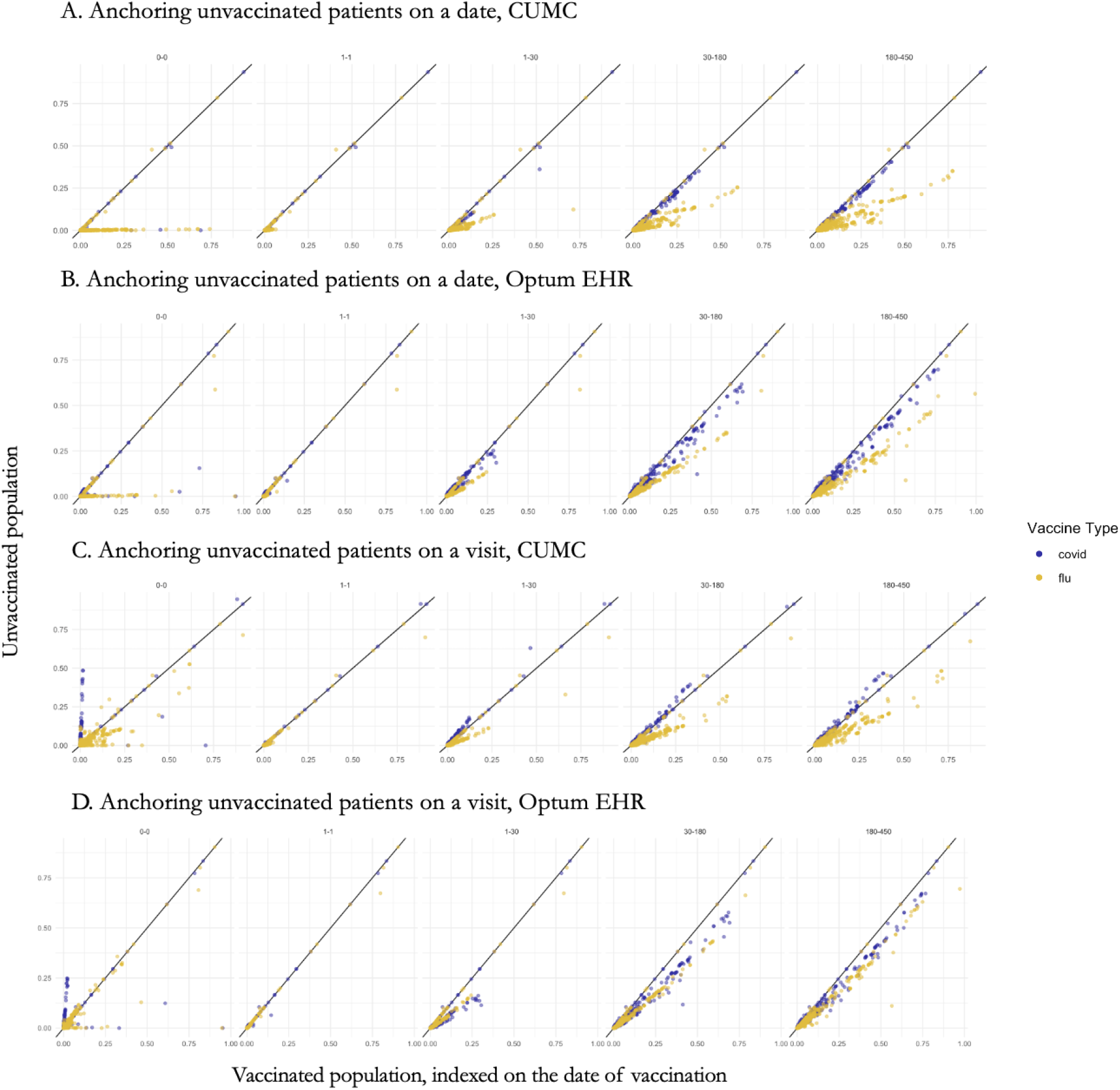
Baseline covariate proportion in vaccinated and unvaccinated populations on day 0, day -1, days -1 to -30, -31 to -180 and -181 to -450 in CUMC and Optum EHR. Each dot represents a covariate; blue – covariate proportion in COVID-19 vaccinated population versus unvaccinated population and yellow – in influenza vaccinated population versus unvaccinated population.

In contrast, comparison with unvaccinated population indexed on a visit (Figure 2, C and D) showed a smaller difference between covariate proportions in CUMC and almost no difference in Optum EHR, potentially indicating that a visit is a better counterfactual for a vaccination date than an arbitrary date.

Covariate proportions in vaccinated patients were closer to the proportions in the unvaccinated population indexed on a visit even with a longer lookback period.

### COVID-19 vaccinated population

As opposed to the influenza vaccinated population, the difference in covariate proportion between COVID-19 vaccinated population and unvaccinated population indexed on an arbitrary date was moderate. We observed that COVID-19 vaccination was associated with a visit in 2.7% of patients (compared to 1.2% on an arbitrary date). In contrast, 55% of influenza vaccinated population had a visit on the date of vaccination (compared to 0.5% of unvaccinated population on an arbitrary date).

Vaccinated population tend to have higher proportion of covariates prior to the index date (looking back a year prior).

When compared to the unvaccinated population indexed on a visit, COVID-19 vaccinated population had markedly lower proportion of most covariates. Those vaccinated with COVID-19 vaccine had much lower rates of diagnoses of both chronic and acute diseases on the date of vaccination compared to a visit in unvaccinated population. The list of conditions included common chronic conditions such as hypertension, depressive disorder, asthma, and diabetes mellitus along with acute conditions like dyspnea, chest pain and fever. Such a difference points out that an arbitrary date may be a better counterfactual for a vaccination date in COVID-19 vaccinated patients.

Comparison of vaccinated patients indexed on the date of vaccination and the same patients indexed on a prior date or visit

### Influenza vaccinated population

Here we compared vaccinated patients indexed on the vaccination date to the same patients indexed on a date or visit within a year prior as we would do in a self-controlled study. We observed that the date of influenza vaccination tended to have a higher proportion of covariates compared to an arbitrary date within a year prior (Figure 3, first column) and even higher compared to an arbitrary visit within a year prior. Patients indexed on the date of vaccination were more likely to have antecedent healthcare encounters, conditions and laboratory tests within a year prior to the vaccination date than within a year prior to their previous visits (Figure 3, C and D). For comparison with an arbitrary date, we observed a mixed effect: in Optum EHR, vaccinated patients had more events preceding their vaccination while in CUMC they had fewer events. Nevertheless, in both data sources the difference between covariate proportions was larger in magnitude when compared to an arbitrary date than when compared to an arbitrary visit.

**Figure 3.**
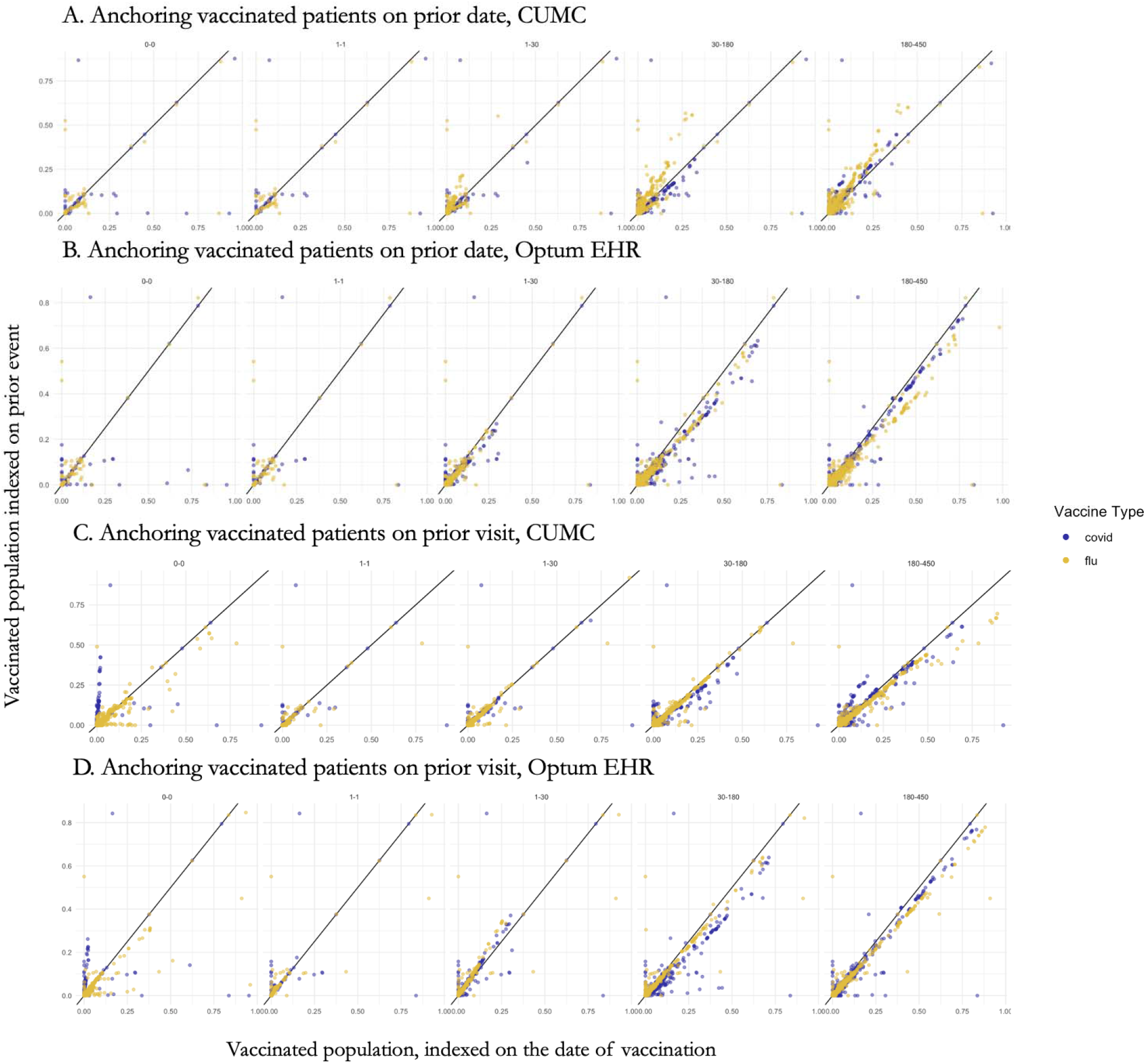
Baseline covariate proportion in vaccinated population indexed on the date of vaccination compared to the same population indexed on a prior visit or date on day 0, day -1, days -1 to -30, -31 to -180 and -181 to -450 in CUMC and Optum EHR. Each dot represents a covariate; blue – covariate proportion in COVID-19 population and yellow – in influenza vaccinated population.

### COVID-19 vaccinated population

The COVID-19 vaccinated population showed a markedly lower proportion of covariates on the day of vaccination compared to a visit or an arbitrary date within a year prior to vaccination. The difference was attenuated with a longer lookback period: COVID-19 vaccinated patients had fewer healthcare events within a year prior to their vaccination compared to their previous history. The difference was less pronounced when compared to an arbitrary date, which, again, points out that an arbitrary date is a better counterfactual for COVID-19 vaccination than an arbitrary visit.

## Discussion

We find that COVID-19 vaccination and influenza vaccination differ drastically from each other, with the proportion of most covariates much higher on the date of vaccination in the influenza group than the COVID-19 group. The results from looking back 31 to 180 days and 181 to 450 days before the vaccination (or index date) may be related to differences in the populations. Persons vaccinated for influenza appear to have more comorbidities and past procedures and measurements than the average population, even after adjusting for age and sex, and persons vaccinated for COVID-19 appear to have a lower proportion of most medical covariates than the average population after adjusting for age and sex. This may be explained if influenza vaccination is targeted to sicker populations on average and if COVID-19 is targeted to the general public, which is healthier on average than those in our electronic health records [8,10].

The drastic effects on day 0—i.e., the day of vaccination and its comparison—are likely related to the context in which the vaccination is given. If the comparison is an arbitrary date in the person’s record, then influenza vaccination has markedly higher covariate proportions, reflecting the association of the vaccination with a healthcare encounter. Moreover, such a trend (not observed for COVID-19 vaccine) was present even when comparing the date of influenza vaccination to the prior patient visits.

The first implication of these results is that, when comparing vaccinated to unvaccinated patients or time, the anchoring event for unvaccinated comparator must be selected carefully. Previous research acknowledged that comparing unexposed and exposed patients in the context of vaccine safety and effectiveness surveillance may lead to between-person confounding due to non-comparable groups [14]. For example, as noted before for influenza, vaccinated and unvaccinated patients differ in co-morbidity prevalence [15]. Nevertheless, even in the same population, the choice of the index date or event influences both baseline covariates and incidence rates of conditions following the index date. For COVID-19 vaccination, it appears that the comparison should not be purposely anchored on a health care visit unless it is a relevant vaccination subgroup (e.g., those vaccinated at hospital discharge).

Adjusting for confounding will be extremely important, as it appears unlikely that a comparison can be chosen perfectly, although the comparisons between the same participants looking a year prior led to the best equivalency for both influenza and COVID-19. Moreover, the difference in patient characteristics require robust selection of covariates for a propensity score model or outcome model as opposed to the current exposed vs unexposed COVID-19 vaccine cohort studies, which only use a limited subset of covariates in their propensity score model [16].

Alternatively, this may argue for a self-controlled study design [17], which mitigates the difference in patient characteristics. However, this design is also sensitive to anchoring (which is what happens on day 0 and around it) and carries other challenges such as accounting for differences in COVID-19 risk over time.

This study has implications beyond using covariates for confounding adjustment. The day 0 results have direct implications for analyses of acute side effects like anaphylaxis that include day 0 because the side effect often occurs immediately. Any study of such short-term effects must directly account for anchoring to the context in which the vaccination is given. Furthermore, studies that compare effectiveness or safety among vaccines must account for differences in populations and in vaccination context. For example, single-dose vaccines may be given preferentially to sicker patients who are unable to return for a second dose, such as those being discharged from the hospital.

## Conclusions

Patient baseline covariates in the unexposed group or time are extremely sensitive to the choice of the index date (anchoring). COVID-19 vaccination and influenza vaccination differ drastically from each other in terms of populations vaccinated and their status on the day of vaccination. Study designs previously used to assess influenza vaccination must be reassessed for COVID-19 to account for a potentially healthier population and lack of medical activity on the day of vaccination.

## Data Availability

All data produced in the present study are available upon reasonable request to the authors

## Ethical approval

The protocol for this research was approved by the Columbia University Institutional Review Board (AAAO7805).

